# Subtle cognitive impairments in memory, attention, and executive functioning in patients with post-COVID syndrome and their relationships with clinical variables and subjective complaints

**DOI:** 10.1101/2022.05.23.22275442

**Authors:** V. Kozik, P. Reuken, I. Utech, J. Gramlich, Z. Stallmach, N. Demeyere, F. Rakers, M. Schwab, A. Stallmach, K. Finke

## Abstract

**Background and objectives:** Cognitive symptoms persisting beyond three months following COVID-19 present a considerable disease burden. We aimed to establish a domain-specific cognitive profile of post-COVID syndrome (PCS) and relationships with subjective cognitive complaints and clinical variables to provide relevant information for the understanding of cognitive dysfunction and its predictors in a clinical cohort with PCS.

**Methods:** In this cross-sectional study, we compared cognitive performance on the clinically viable Oxford Cognitive Screen-Plus between a large post-COVID cohort (*n* = 282) and a socio-demographically matched healthy control group (*n* = 52). We assessed group differences in terms of fatigue and depression as well as relationships between cognitive dysfunction and clinical and patient-reported outcomes.

**Results:** On a group-level, patients scored significantly lower on delayed verbal memory (non-parametric effect size *r* = .13), attention (*r* = .1), and executive functioning (*r*=.1) than healthy controls. In each of these domains, 10-20% of patients performed more than 1.5 SD below the healthy control mean. Delayed Memory was particularly affected and a small proportion of its variance was explained by hospitalisation (*β* = -.72, *p* < .01) and age (*β* = -.03, *p* < .05; R^2^adj. = .08). Attention scores were significantly predicted by hospitalisation (*β* = -.78, *p* < .01) and fatigue (*β* = -.04, *p* < .05; R^2^adj. = .06).

**Discussion:** PCS is associated with long-term cognitive dysfunction, particularly in delayed verbal memory, attention, and executive functioning. Deficits in delayed memory performance seem to be of particular relevance to patients’ subjective experience of impairment. Initial disease severity, current level of fatigue, and age seem to predict cognitive performance, while time since infection, depression, and pre-existing conditions do not. Longitudinal data are needed to map long-term course of cognitive dysfunction in PCS.

## 1 Introduction

A considerable number of individuals affected by coronavirus disease 2019 (COVID-19), including mild and asymptomatic cases, report long-term cognitive effects, in addition to fatigue and physical symptoms (e.g., ^1-5^; for reviews see ^6,7^). If symptoms develop during or after infection, persist for more than 12 weeks and cannot be explained by another diagnosis, the patient suffers from post-COVID-19 syndrome (PCS), according to NICE guidelines^8^. A recent study with 355 patients from a post-COVID outpatient clinic^a^, reported that over 90% reported signs of fatigue and depression and 23% performed below cut-off in a cognitive screening (Montreal Cognitive Assessment; MoCA^9^). Similar incidences of below cut-off MoCA scores were reported following SARS-CoV-2 infection in a recent population-based study^10^.

Long-term cognitive deficits are of high clinical relevance, as they have a strong impact on patients’ daily functioning, employment, and the ability to return to work, and thus, constitute a large disease burden^11^. A characterisation of the cognitive profile in PCS and the relationships of deficits in different domains with subjective cognitive complaints and relevant clinical variables is of the essence, as it could foster the understanding of the underlying pathogenic mechanisms and improve knowledge of the course of the syndrome. However, the overall cut-offs for short cognitive screens are not suitable for such analyses.

Initial evidence from studies using more comprehensive test batteries point towards deficits in the domains of attention, memory, and executive functioning following SARS-CoV-2 infection (e.g., ^12-15^, for review see ^16^). However, in these studies, samples were either small or assessed remotely in uncontrolled settings, participants did not consistently meet criteria for the diagnosis of initial infection or PCS^8^, and/or healthy control groups were missing.

For the reliable identification of a domain-specific neuropsychological profile and the clinical factors influencing the domains, it is crucial to assess large, well-defined patient groups with appropriate assessment tools. Furthermore, comparisons with matched, healthy groups are needed to control for the potential influence of generally increased psychological stress under conditions of a pandemic on cognitive functions. However, the use of comprehensive neuropsychological batteries, particularly in a standardised, in-person setting is not easily scalable, as it is time-consuming regarding application, scoring, and interpretation, and requires specialised staff.

The present study used a clinically suitable, time- and cost-effective alternative to meet these challenges. The Oxford Cognitive Screen-Plus^b^ (OCS-Plus^17^) is a tablet-based screening tool, which bridges the gap between short-from screens and comprehensive neuropsychological batteries, in terms of resource-efficiency and its psychometric properties. It facilitates a more detailed screening of domain-specific cognitive functions and the establishment of a profile of spared and affected domains in subclinical and clinical populations^17,18^. Its use requires little training from operating staff and outcome measures are scored automatically.

Using this innovative test, the first aim of this study was to elucidate the cognitive profile associated with PCS by assessing all potentially relevant domains with a large, clinical sample in comparison to a healthy control group, matched by age, sex, and education. The second aim was to establish relationships between affected cognitive domains and subjective cognitive complaints, as well as relevant clinical variables, such as initial disease severity, time since infection, age, depression, fatigue, and comorbidities in order to identify predictors of specific cognitive deficits in this clinically referred, well-defined post-COVID cohort.

## 2 Methods

### 2.1 Participants

A total of 282 patients and 52 healthy controls were included in this study. We included all patients who presented to the post-COVID outpatient clinic at Jena University Hospital (Germany) between August 2020 and March 2022 and who had previously been confirmed positive for SARS-CoV-2 using a PCR-test, were willing and able to give informed consent, and were capable of taking part in the assessment. We further only included participants in either group, who did not have a history of relevant neurological or severe psychiatric disorders potentially impairing cognition, substance use disorder, or relevant vision and hearing problems, and who were between the ages of 18 and 65. We chose the upper age limit to avoid any issues pertaining to cognitive changes due to age-associated neurodegenerative processes. Of 399 patients, who initially presented to the clinic within the given time period, met inclusion criteria and consented to their participation, data for 76 patients is not available due to either technical difficulties before or during testing, data for 39 patients is unavailable due to logistical issues or constraints in the clinical setting, and two participants withdrew consent after testing. For a patient-only regression analysis with six predictor variables, we have 80% power to detect effects larger than R^2^ = .05 with our smallest sub-sample (alpha = .05). Based on the fact of relatively low variability and near ceiling performance on the relevant domain scores of the OCS-Plus by healthy, largely elderly participants (see table 8 in ^17^), we expect our smaller, but socio-demographically matched control group to strike the balance between sufficiency to represent healthy variability and resource efficiency.

### 2.2 Assessment

Patients underwent structured anamnesis including the patient’s medical history, basic socio-demographic data, and subjective cognitive complaints. All participants completed the depression module of the Patient Health Questionnaire (PHQ-9^19^), the Fatigue Assessment Scale (FAS^20^) and the Brief Fatigue Inventory (BFI^21^). In the same session, cognitive functioning was assessed by research staff using the Oxford Cognitive Screen-Plus^17^, which consists of 9^c^ subtasks: Picture Naming, Semantics, Orientation, Word Memory Encoding, Delayed Recall, Trails, Episodic Recognition, Figure Copy, and Cancellation (see ^17^ and table 1).

**Table 1:**
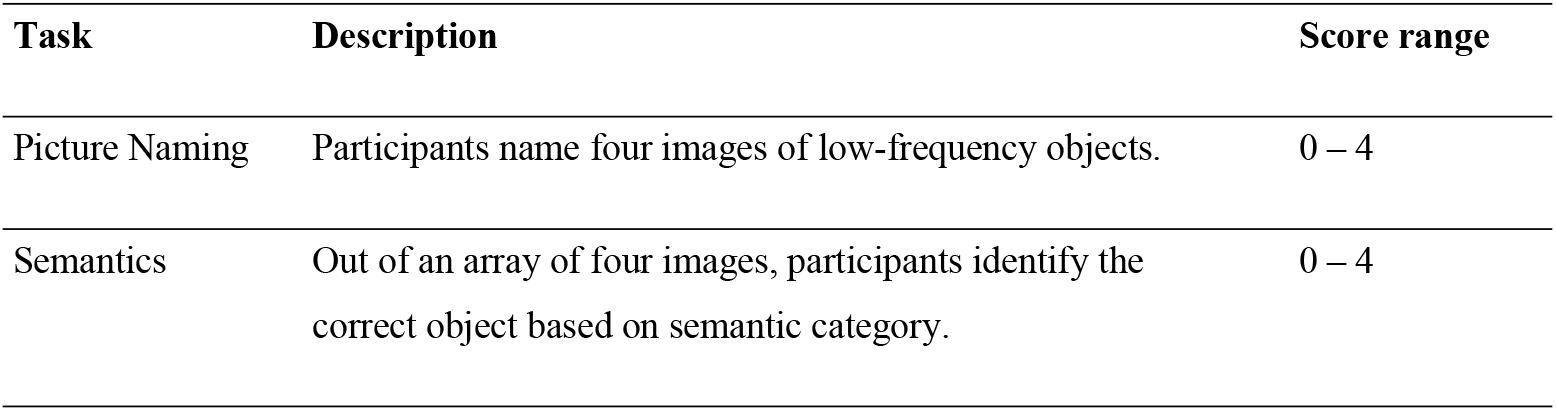
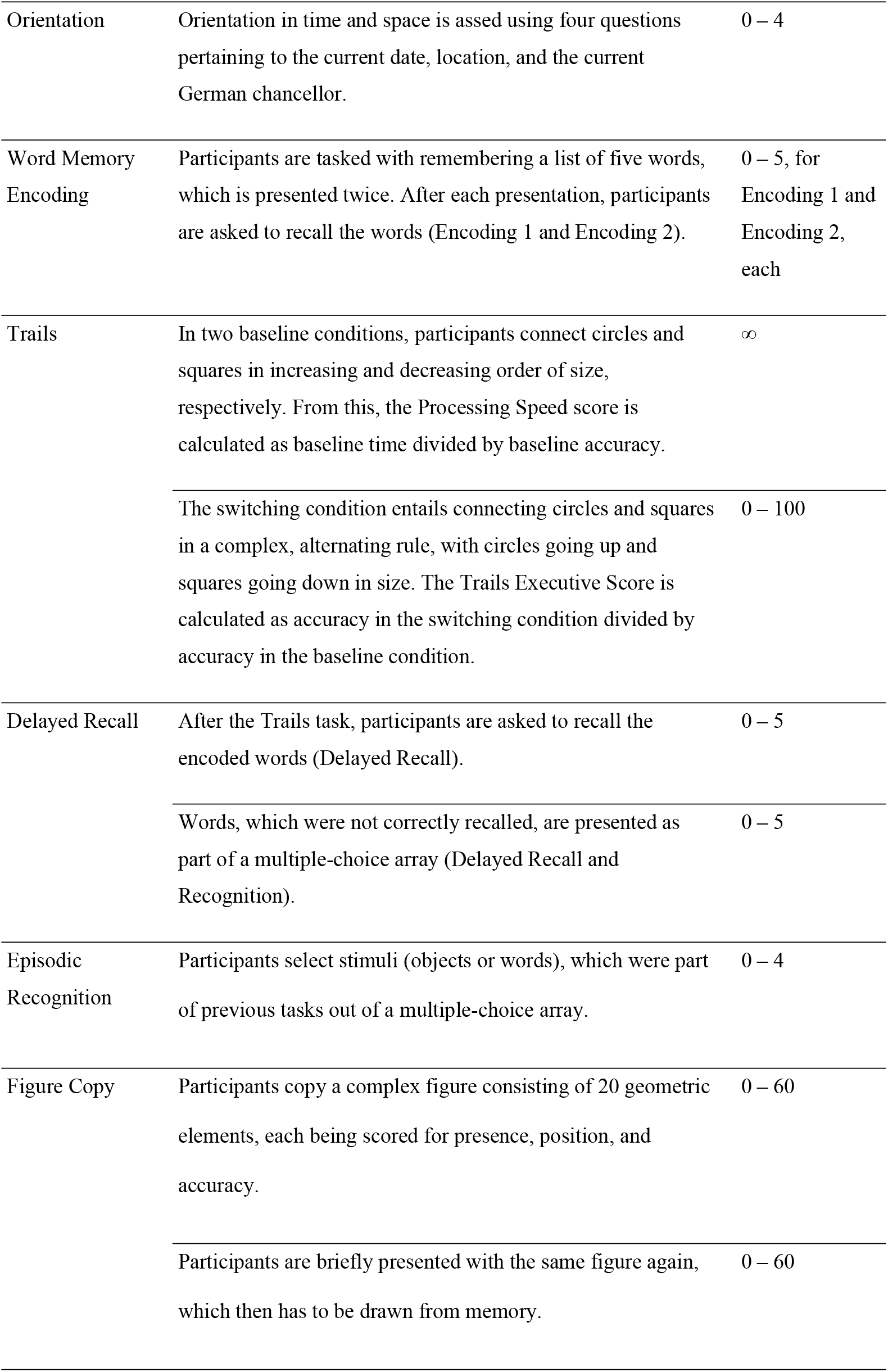
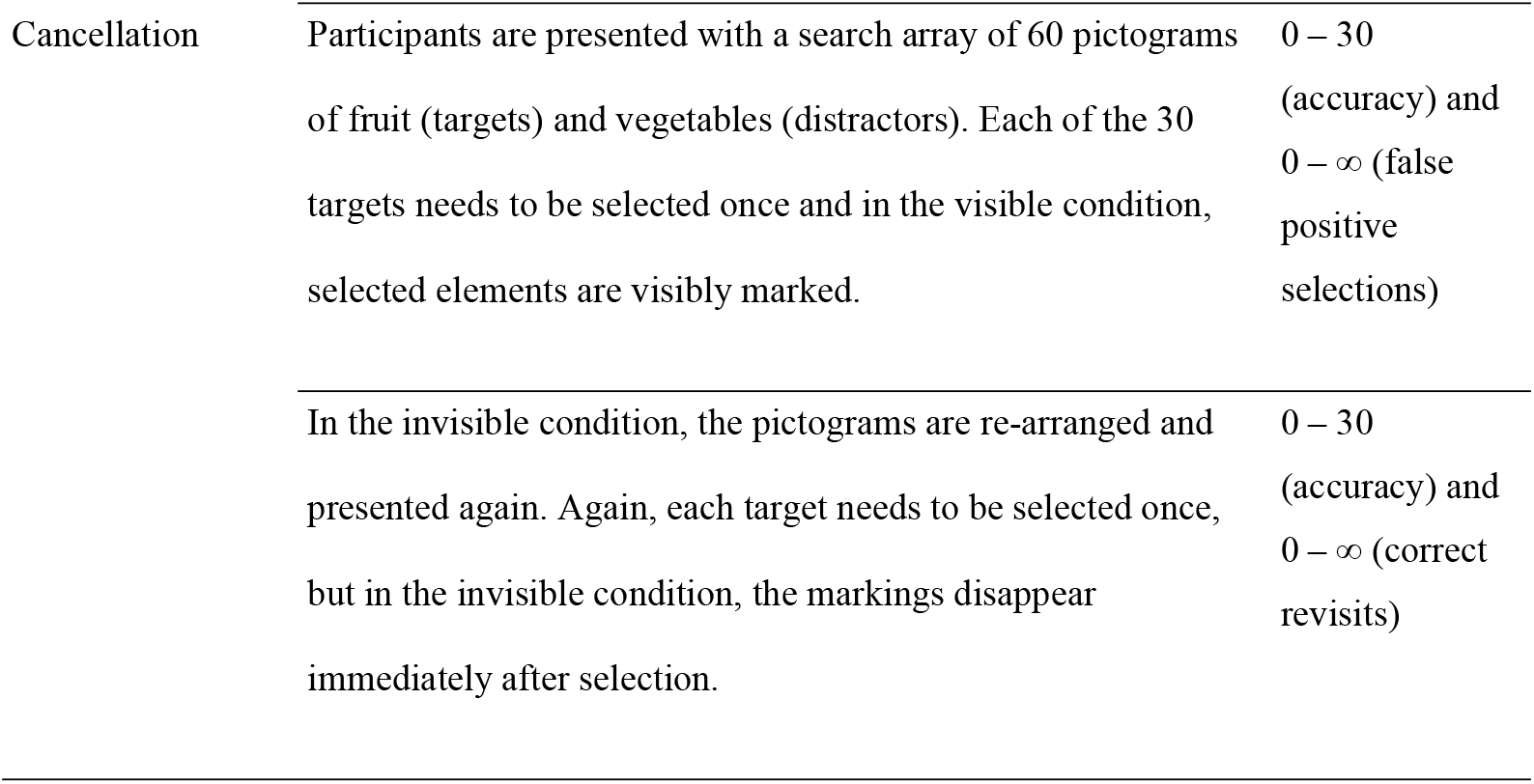
OCS-Plus tasks descriptions

| Task | Description | Score range |
| --- | --- | --- |
| Picture Naming | Participants name four images of low-frequency objects. | 0 – 4 |
| Semantics | Out of an array of four images, participants identify the correct object based on semantic category. | 0 – 4 |

<sup>c</sup> Due to time constraints in the clinical setting, the OCS-Plus subtask “rule finding” was skipped. Accordingly, scoring for executive functioning differs from the method proposed by Demeyere et al. (2021).

|  |  |  |
| --- | --- | --- |
| Orientation | Orientation in time and space is assessed using four questions pertaining to the current date, location, and the current German chancellor. | 0 – 4 |
| Word Memory Encoding | Participants are tasked with remembering a list of five words, which is presented twice. After each presentation, participants are asked to recall the words (Encoding 1 and Encoding 2). | 0 – 5, for Encoding 1 and Encoding 2, each |
| Trails | In two baseline conditions, participants connect circles and squares in increasing and decreasing order of size, respectively. From this, the Processing Speed score is calculated as baseline time divided by baseline accuracy. | ∞ |
|  | The switching condition entails connecting circles and squares in a complex, alternating rule, with circles going up and squares going down in size. The Trails Executive Score is calculated as accuracy in the switching condition divided by accuracy in the baseline condition. | 0 – 100 |
| Delayed Recall | After the Trails task, participants are asked to recall the encoded words (Delayed Recall). | 0 – 5 |
|  | Words, which were not correctly recalled, are presented as part of a multiple-choice array (Delayed Recall and Recognition). | 0 – 5 |
| Episodic Recognition | Participants select stimuli (objects or words), which were part of previous tasks out of a multiple-choice array. | 0 – 4 |
| Figure Copy | Participants copy a complex figure consisting of 20 geometric elements, each being scored for presence, position, and accuracy. | 0 – 60 |
|  | Participants are briefly presented with the same figure again, which then has to be drawn from memory. | 0 – 60 |
| Cancellation | Participants are presented with a search array of 60 pictograms of fruit (targets) and vegetables (distractors). Each of the 30 targets needs to be selected once and in the visible condition, selected elements are visibly marked. | 0 – 30 (accuracy) and 0 – ∞ (false positive selections) |
| | In the invisible condition, the pictograms are re-arranged and presented again. Again, each target needs to be selected once, but in the invisible condition, the markings disappear immediately after selection. | 0 – 30 (accuracy) and 0 – ∞ (correct revisits) |

Assessment takes approximately 25 minutes and is completed using a stylus pen on a tablet computer. From the OCS-Plus subtasks, six domain scores may be calculated: Naming and Semantic Understanding (Picture Naming + Semantics), Memory Encoding (Encoding 1 + Encoding 2), Delayed Memory (Delayed Recall + Delayed Recall and Recognition), Praxis (Figure Copy + Figure Recall), Attention (Cancellation + Invisible Cancellation), and Executive Functioning (Trails Executive Score – Cancellation false positives^17^).

### 2.3 Statistical Analysis

We compared socio-demographic variables between patients and healthy controls using t-tests with Welch correction to account for the difference in sample sizes and a chi-squared test with Yates’ continuity correction to compare sex ratios. Fatigue and depression scores were compared between groups using Welch two sample t-tests. Performance on the OCS-Plus subtasks and overall domain scales was compared between patients and controls using Wilcoxon rank sum tests with continuity correction. Based on the current state of the literature, we selected subtasks and overall domain scores as particularly relevant to our analyses, which capture the domains of attention, memory, and executive functioning. As we expected patients to perform worse in these domains, we used one-tailed tests (alpha = .05). We then examined how many patients fell below a cut-off of 1.5 standard deviations below the healthy sample means on the domains of interest. To take into account the heterogeneity of symptoms and particularly of cognitive complaints, we split patients into two groups: those who complain of both memory and concentration problems (high complainers) and those who report only one or none of these symptoms (low complainers). These groups were then compared in terms of their cognitive performance on the domain scales of the OCS-Plus. To explore predictors of attentional, memory, and executive functioning problems as part of post-COVID syndrome, we performed multiple linear regression analyses within the patient cohort. We included initial disease severity, age, days since infection, fatigue, and relevant comorbidities as predictors of performance on the OCS-Plus. The need for hospitalisation i.e., outpatient versus inpatients treatment, was used as a proxy for initial disease severity. Comorbidities were included as an index of five binarised pre-existing conditions: hypertension, coronary heart disease, chronic heart failure, diabetes mellitus, and psychiatric disorders (range: 0 – 5). To estimate generalisability of the models, we computed nonparametric bootstrap (2,000 replications) confidence intervals around coefficients. We then separated patients into two groups for each pre-existing condition, i.e., condition “present” and “not present”, and compared groups on each cognitive domain to assess the effect of the individual conditions. We corrected for multiple comparisons using the Benjamini-Hochberg procedure (Q = 5%). For each OCS-Plus score, an average 0.12% of data points are missing due to technical difficulties. Analysis was performed using R version 4.2.0^22^.

### 2.4 Standard Protocol Approvals, Registrations, and Patient Consents

Written informed consent was obtained from all participants. The study was approved by the ethics committee of the Jena University Hospital [amendment to 5082-02/17].

### 2.5 Data Availability

Anonymised data not published in this article will be made available upon reasonable request.

## 3 Results

### 3.1 Sociodemographic and clinical description of post-COVID-19 patients and healthy controls

Basic socio-demographic information for both groups are presented in table 2. There were no differences between groups in terms of age (*t* = -.76, *p* = .451), education (*t* = 1.72, *p* = .09), or sex ratios (chi-squared = .61, *p* = .435). Please refer to table 3 for an overview of patient clinical data stratified by hospitalisation.

**Table 2:** Socio-demographic distribution by group

|  | Controls (n = 52) | Patients (n = 282) | Total (N = 334) |
| --- | --- | --- | --- |
| **Age** |  |  |  |
| Mean (SD) | 45.62 (10.15) | 46.80 (11.27) | 46.61 (11.15) |
| Range | 22.00 – 65.00 | 18.00 – 65.00 | 18.00 – 65.00 |
| **Sex** |  |  |  |
| Female | 31 (59.6%) | 186 (66.0%) | 217 (65.0%) |
| Male | 21 (40.4%) | 96 (34.0%) | 117 (35.0%) |
| **Education (years)** |  |  |  |
| Missing (n) | 0 | 28 | 28 |
| Mean (SD) | 15.28 (1.97) | 14.76 (2.07) | 14.85 (2.06) |
| Range | 11.00 – 18.00 | 10.00 – 18.00 | 10.00 – 18.00 |

Note. SD = standard deviation

**Table 3:** Clinical data stratified by need for hospitalisation

| Clinical Variable | Distribution | Missingness |
| --- | --- | --- |
| Weeks since infection, M(SD, Range) | 37.3 (17.6, 12 – 104) | 0 |
| **Outpatient treatment, n(%)** | 215 (76.2) | 0 |
| WHO severity grade, n(%) |  | 1 |
| 1 | 3 (1.4) |  |
| 2 | 210 (98.1) |  |
| 3 | 1 (0.5) |  |
| **Comorbidities** |  | 0 |
| Cardiovascular diseases, n(%) | 60 (27.9) |  |
| Diabetes mellitus, n(%) | 4 (1.9) |  |
| Psychiatric comorbidities, n(%) | 29 (13.5) |  |
| **Inpatient treatment, n(%)** | 67 (23.8) | 0 |
| Hospital stay (days), M(SD) | 12 (8.8) | 0 |
| Oxygen support, n(%) | 48 (71.6) | 0 |
| ICU admission, n(%) | 22 (33.3) | 1 |
| ICU stay (days), M(SD) | 3.2 (7.8) | 4 |
| WHO severity grade, n(%) |  | 0 |
| 2 | 4 (6.3) |  |
| 3 | 11 (16.4) |  |
| 4 | 26 (38.8) |  |
| 5 | 20 (29.9) |  |
| 7 | 6 (9.0) |  |
| Comorbidities |  | 0 |
| Cardiovascular diseases, n(%) | 43 (64.2) |  |
| Diabetes mellitus, n(%) | 10 (14.9) |  |
| Psychiatric comorbidities, n(%) | 11 (16.4) |  |

*Note.* M = mean; SD = standard deviation; WHO = World Health Organization; ICU = intensive care unit; cardiovascular diseases included hypertension, coronary heart disease, and chronic heart failure

### 3.2 Subjective cognitive complains, depression, and fatigue

During the anamnestic interview, 69.9% of patients complained of attention and 58.9% of memory problems. 55.7% of patients complained of both attention and memory problems. As the two fatigue questionnaires were highly correlated (*r*(329) = .78, *p* < .0001), only the results from the FAS will be used for further analysis. Patients scored significantly higher on the FAS (*r* = .50, *p* < .0001) and on the PHQ-9 (*r* = .44, *p* < .0001) than controls (see table 4 for complete results).

**Table 4:** Questionnaire data for controls and patients

| Questionnaire | Controls |  |  |  | Patients |  |  |  | Wilcoxon rank sum test |  |
| --- | --- | --- | --- | --- | --- | --- | --- | --- | --- | --- |
|  | n | Mean | SD | SEM | n | Mean | SD | SEM | *W* | *r*<sup>a</sup> |
| FAS | 50 | 17.30 | 4.81 | 0.68 | 282 | 31.27 | 9.05 | 0.54 | 1360 | 0.50*** |
| PHQ-9 | 50 | 3.92 | 2.93 | 0.41 | 282 | 10.69 | 5.57 | 0.33 | 2004.5 | 0.44*** |

*Note.* FAS = Fatigue Assessment Scale; PHQ-9 = Patient Health Questionnaire; SD = standard deviation; SEM = standard error of the mean; *r* = effect size; <sup>a</sup> \* *p*<.5; \*\**p*<.01; \*\*\**p*<.001

### 3.3 Comparison between patients and healthy controls on OCS-Plus subtasks

Patients scored lower than healthy controls on the tasks Encoding 2 (*r* = .1, *p* = .034), Delayed Recall accuracy (*r* = .12, *p* = .013), Figure Copy accuracy (*r* = .1, *p* = .037), Cancellation false positives (*r* = -.1, *p* = .03), and Invisible Cancellation accuracy (*r* = .12, *p* = .018). However, none of the comparisons survived correction. Please refer to table 5 for full results.

**Table 5:** Performance on the OCS-Plus subtasks per group

| OCS-Plus task | Controls |  |  |  |  | Patients |  |  |  |  | Wilcoxon rank sum test |  |  |
| --- | --- | --- | --- | --- | --- | --- | --- | --- | --- | --- | --- | --- | --- |
|  | n | Mean | Median | SD | SEM | n | Mean | Median | SD | SEM | *W* | *p* | *r* |
| Picture Naming accuracy | 52 | 3.96 | 4.00 | 0.19 | 0.03 | 280 | 3.95 | 4.00 | 0.26 | 0.02 | 7340 | 0.397 |  |
| Semantics accuracy | 51 | 3.92 | 4.00 | 0.27 | 0.04 | 282 | 3.89 | 4.00 | 0.33 | 0.02 | 7419.5 | 0.249 |  |
| Orientation accuracy | 52 | 3.90 | 4.00 | 0.30 | 0.04 | 282 | 3.96 | 4.00 | 0.19 | 0.01 | 6913 | 0.962 |  |
| *Encoding 1* | 52 | 4.54 | 5.00 | 0.64 | 0.09 | 274 | 4.52 | 5.00 | 0.66 | 0.04 | 7158.5 | 0.475 |  |
| *Encoding 2* | 52 | 5.00 | 5.00 | 0.00 | 0.00 | 279 | 4.94 | 5.00 | 0.24 | 0.01 | 7696 | 0.034 | 0.1* |
| *Delayed Recall accuracy* | 50 | 3.78 | 4.00 | 1.13 | 0.16 | 279 | 3.24 | 3.00 | 1.48 | 0.09 | 8327 | 0.013 | 0.12* |
| *Delayed Recall and Recognition* | 51 | 4.88 | 5.00 | 0.33 | 0.05 | 279 | 4.74 | 5.00 | 0.56 | 0.03 | 7818 | 0.052 |  |
| *Episodic Recognition accuracy* | 51 | 3.57 | 4.00 | 0.57 | 0.08 | 280 | 3.42 | 4.00 | 0.72 | 0.04 | 7768 | 0.13 |  |
| *Trails Executive Score* | 51 | 88.54 | 100.00 | 16.30 | 2.28 | 278 | 82.03 | 92.86 | 24.31 | 1.46 | 7918 | 0.078 |  |
| *Processing Speed* | 51 | 16.62 | 14.64 | 7.06 | 0.99 | 278 | 15.03 | 13.39 | 6.91 | 0.41 | 8094 | 0.054 |  |
| Figure Copy accuracy | 43 | 56.98 | 58.00 | 2.88 | 0.44 | 279 | 56.39 | 57.00 | 2.85 | 0.17 | 7002.5 | 0.037 | 0.1* |
| Figure Recall accuracy | 42 | 47.12 | 47.50 | 7.98 | 1.23 | 279 | 46.53 | 48.00 | 8.43 | 0.50 | 6022 | 0.386 |  |
| *Cancellation accuracy* | 51 | 29.80 | 30.00 | 0.45 | 0.06 | 275 | 29.77 | 30.00 | 0.52 | 0.03 | 7162 | 0.363 |  |
| *Cancellation false positives* | 51 | 0.04 | 0.00 | 0.20 | 0.03 | 276 | 0.16 | 0.00 | 0.44 | 0.03 | 6390 | 0.03 | -0.1* |
| *Invisible Cancellation accuracy* | 51 | 29.02 | 29.00 | 0.97 | 0.14 | 275 | 28.45 | 29.00 | 1.55 | 0.09 | 8267 | 0.018 | 0.12* |
| *Invisible Cancellation correct revisits* | 52 | 1.08 | 0.50 | 1.38 | 0.19 | 276 | 1.43 | 1.00 | 1.91 | 0.11 | 6570 | 0.155 |  |

*Note.* OCS-Plus = Oxford Cognitive Screen-Plus; SD = standard deviation; SEM = standard error of the mean; *r* = effect size; \* = significant before FDR-correction, \*\* = significant after FDR-correction; italicised: subtasks of interest

### 3.4 Comparison between patients and healthy controls on OCS-Plus domain scales

Patients scored significantly lower than healthy controls on the scales of Delayed Memory (*r* = .13, *p* < .01), Executive Functioning (*r* = .1, *p* = .033), and Attention (*r* = .1, *p* = .027). See table 6 and figure 1 for results stratified by group.

**Table 6:** Performance on the OCS-Plus domain scales per group

| OCS-Plus scale | Controls |  |  |  |  | Patients |  |  |  |  | Wilcoxon rank sum test |  |  |
| --- | --- | --- | --- | --- | --- | --- | --- | --- | --- | --- | --- | --- | --- |
|  | n | Mean | Median | SD | SEM | n | Mean | Median | SD | SEM | W | p | r |
| Naming and Semantic Understanding | 51 | 7.88 | 8 | 0.38 | 0.05 | 280 | 7.83 | 8 | 0.44 | 0.03 | 7478 | 0.185 |  |
| *Memory Encoding* | 52 | 9.54 | 10 | 0.64 | 0.09 | 271 | 9.46 | 10 | 0.75 | 0.05 | 7296 | 0.322 |  |
| *Delayed Memory* | 50 | 8.68 | 9 | 1.24 | 0.17 | 279 | 7.98 | 8 | 1.77 | 0.11 | 8438.5 | <0.01 | 0.13** |
| Praxis | 43 | 103.86 | 105 | 8.92 | 1.36 | 279 | 102.92 | 105 | 9.63 | 0.58 | 6189 | 0.369 |  |
| *Executive Functioning* | 50 | 88.7 | 100 | 16.46 | 2.33 | 275 | 81.59 | 92.86 | 24.79 | 1.49 | 7945 | 0.033 | 0.1** |
| *Attention* | 50 | 58.82 | 59 | 1.06 | 0.15 | 275 | 58.22 | 59 | 1.72 | 0.10 | 8021 | 0.027 | 0.1** |

Note. OCS-Plus = Oxford Cognitive Screen-Plus; SD = standard deviation; SEM = standard error of the mean; r = effect size; \* = significant before FDR-correction, \*\* = significant after FDR-correction; italicised: domain scales of interest

**Figure 1:**
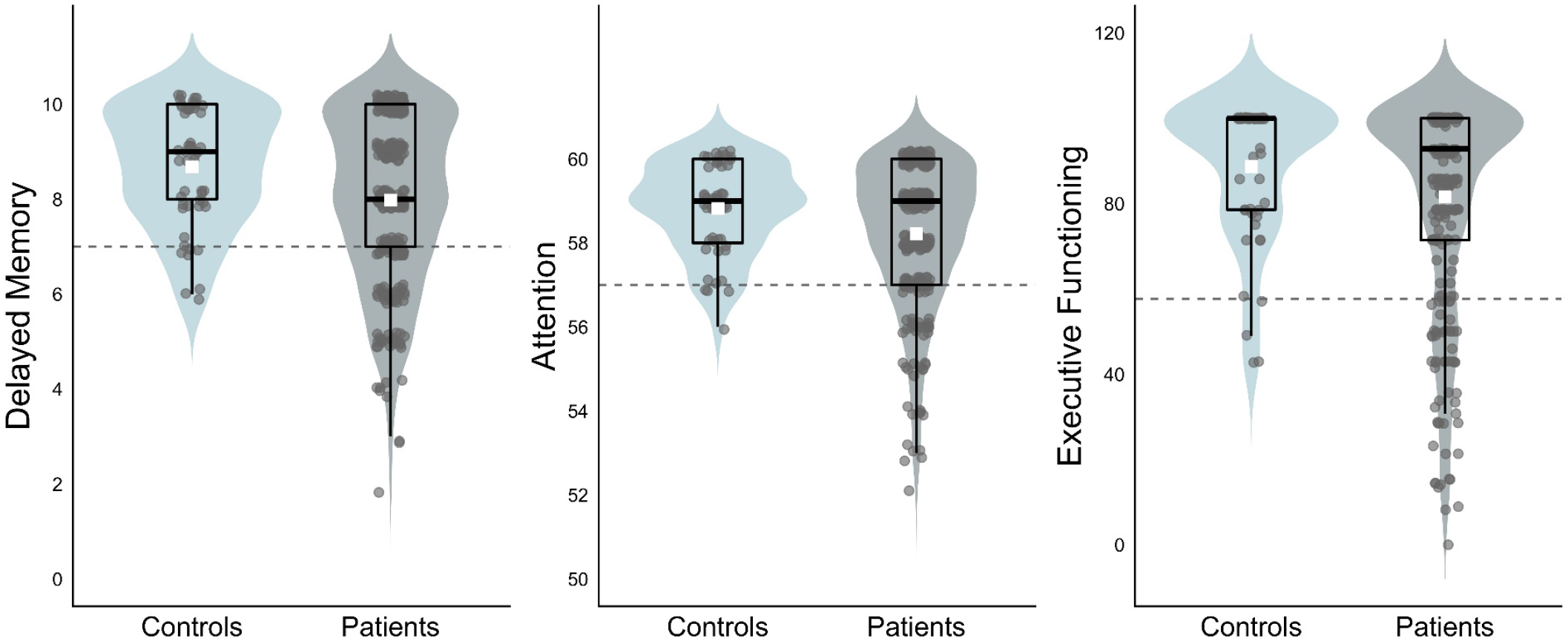
Performance on the OCS-Plus domain scales by controls and patients with post-COVID syndrome *Note*. Distribution of scores on the OCS-Plus domain scales of Delayed Memory, Attention, and Executive Functioning, per group. Grey dots represent individual participants, white squares represent group mean, and dashed lines represent cut-off (1.5 standard deviations below control mean).

### 3.5 Proportional impairment per group on OCS-Plus domain scales

10.7% of patients scored below the cut-off on Memory Encoding (versus 3.85% of controls), 21.15% of patients scored below the cut-off on Delayed Memory (versus 6% of controls), 19.27% of patients scored below the cut-off on Executive Functioning (versus 8% of controls), and 14.91% of patients scored below the cut-off on Attention (versus 2% of controls; see figure 2, panel A). Out of those patients for whom there is complete data for all domain scores, 53.7% of patients were impaired on at least one domain score (versus 25% of controls), 18.68% scored below the cut-off on at least 2 domains (versus 5% of controls), and 3.89% scored below the cut-off on at least 3 domains (versus 0% of controls; see figure 2, panel B).

**Figure 2:**
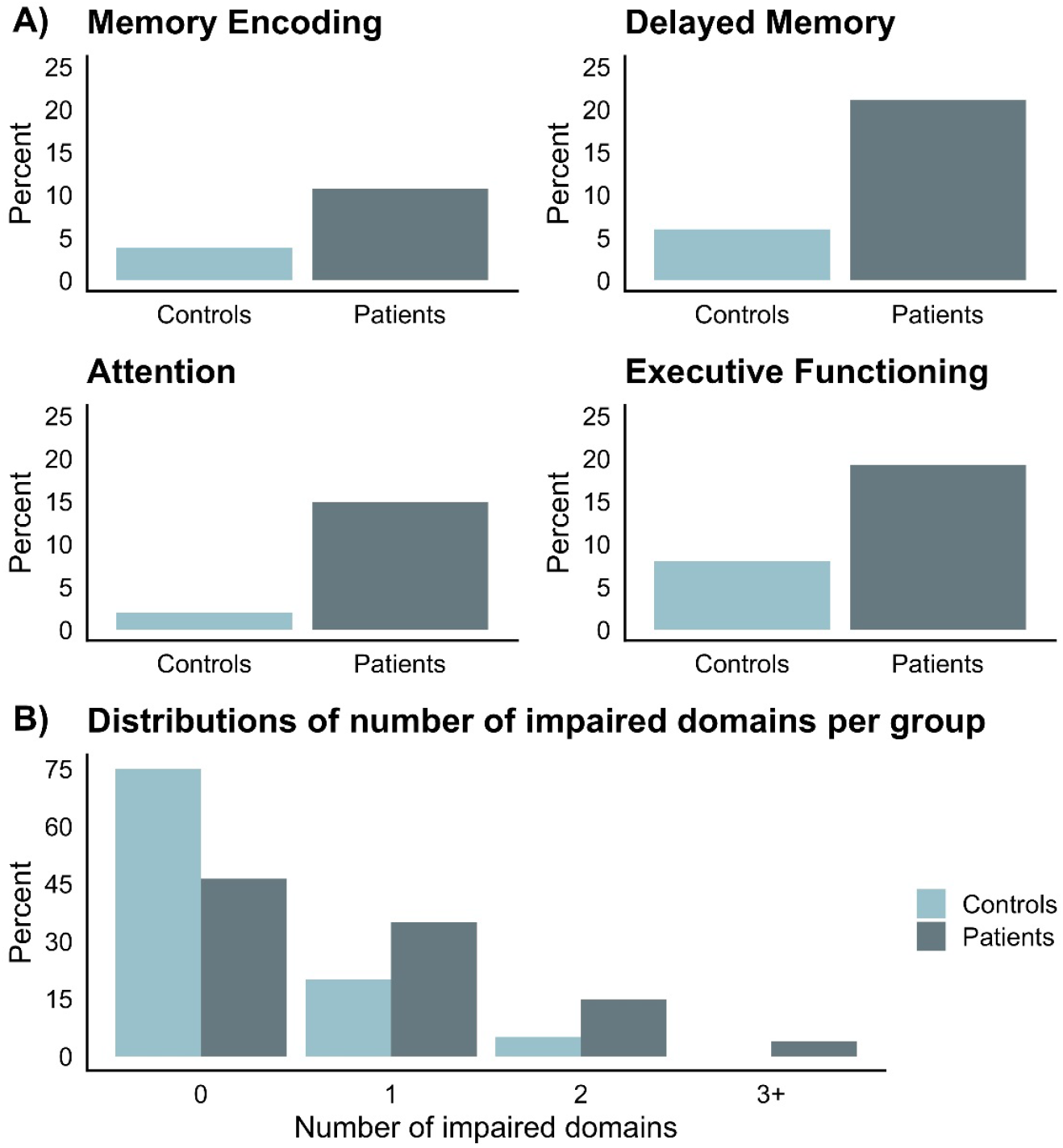
Distributions of patients and controls scoring below cut-off on OCS-Plus domain scores *Note*. Panel A) Percentage of participants under the cut-off (1.5 standard deviations below control mean) per group in the domain scores of interest. Panel B) Percentage of participants per group, who fall under the cut-off in no, one, two, or at least three domain scores.

### 3.6 Relationship between subjective cognitive complaints and Delayed Memory performance

76 patients reported no or only one cognitive symptom and 206 patients reported both attention and memory difficulties. Performance in the Delayed Memory domain differed between those with and those without subjective cognitive complaints (*W* = 8669, *p* = .024). There were no performance differences on any other domain scale (see table S1 in the supplemental material for complete results).

### 3.7 Relationships between clinical variables and performance on the domains of Delayed Memory, Attention, and Executive Functioning

The overall model to predict Delayed Memory performance, with hospitalisation, age, days since infection, fatigue, and comorbidities as predictors was significant (F(6, 272) = 4.84, *p* <.001, R^2^adj.= .08). Hospitalisation (*β* = -.72, *p* < .01) and age (*β* = -.03, *p* <.05) significantly predicted Delayed Memory performance. The model to predict performance on the Attention domain score was also significant (F(6, 268) = 4.07, *p* <.001, R^2^adj.= .06), with hospitalisation (*β* = -.78, *p* <.01) and fatigue (*β* = -.04, *p* <.05) as significant predictors. The model to predict performance in the Executive Functioning domain was not significant. Please see table 7 for full results.

**Table 7:** Coefficient-level estimates for models fitted to estimate variation in (1) Delayed Memory, (2) Attention, and (3) Executive Functioning performance

|  | (1) Delayed Memory | (2) Attention | (3) Executive Functioning |
| --- | --- | --- | --- |
|  | *OLS* | *OLS* | *OLS* |
| Intercept | 9.564***<br>(8.383, 10.789) | 59.748***<br>(58.576, 60.844) | 81.631***<br>(59.821, 99.055) |
| Hospitalisation (inpatient) | -0.724**<br>(-1.205, -0.265) | -0.779**<br>(-1.402, -0.239) | -2.314<br>(-10.856, 5.074) |
| Age | -0.025* | -0.014 | 0.069 |
|  | **(-0.042, -0.005)** | (-0.032, 0.003) | (-0.200, 0.394) |
| Days since infection | 0.002*<br>(0.000, 0.003) | 0.001<br>(0.000, 0.003) | -0.025*<br>(-0.049, -0.004) |
| FAS | -0.018<br>(-0.052, 0.013) | **-0.043\***<br>**(-0.075, -0.012)** | 0.184<br>(-0.357, 0.676) |
| PHQ-9 | -0.009<br>(-0.060, 0.042) | 0.035<br>(-0.011, 0.086) | -0.029<br>(-0.722, 0.767) |
| Comorbidities | -0.031<br>(-0.321, 0.239) | -0.055<br>(-0.340, 0.232) | -2.505<br>(-6.728, 1.762) |
| Observations | 279 | 275 | 275 |
| R<sup>2</sup> | 0.096 | 0.083 | 0.027 |
| Adjusted R<sup>2</sup> | 0.076 | 0.063 | 0.005 |
| Residual Std. Error | 1.702 (df = 272) | 1.668 (df = 268) | 24.727 (df = 268) |
| F Statistic | 4.838*** (df = 6; 272) | 4.067*** (df = 6; 268) | 1.223 (df = 6; 268) |

*Note:* Coefficients and confidence intervals (nonparametric bootstrap, 2,000 replications, in parentheses), bolded: significant estimates, with bootstrap confidence intervals not overlapping zero; \* $p < 0.05$ ; \*\* $p < 0.01$ ; \*\*\* $p < 0.001$

On the level of individual comorbidities, those with hypertension performed worse in the Delayed Memory domain. No other comparisons between groups with and without individual comorbidities survived correction. Regression analysis revealed no effect of hypertension on Delayed Memory performance, when controlling for our set of covariates (see S2-S4 in the supplemental material).

## 4 Discussion

In this study, subtle^d^, but meaningful impairments in attention, delayed memory, and executive functions as well as preserved basic orientation, language, and visuo-spatial functions were identified in patients with post-COVID syndrome (PCS). High levels of patients’ subjective cognitive complaints were associated with poorer performance on the delayed memory scale, but not other cognitive domains. In regression analyses we found significant clinical predictors of memory and attentional performance, but none for executive functions. Specifically, we found that initial disease severity predicted performance in the domains of attention and delayed recall, in the sense that hospitalised patients performed significantly worse than non-hospitalised patients. Further, older age predicted worse performance on the delayed memory domain and higher levels of fatigue predicted worse performance on the domain of attention. We found no associations between delayed memory or attentional performance and time passed since infection, depression, or comorbidities.

The identified neuropsychological profile of patients with PCS fits with results of early studies (e.g., ^12-14^). However, the present study goes beyond these prior studies by documenting persisting deficits in a large patient sample with previous SARS-CoV-2 infection confirmed by laboratory testing and fulfilling the NICE criterion of symptom persistence beyond 12 weeks post-infection^8^ in comparison to a socio-demographically matched control group. Moreover, our participants were assessed in a face-to-face setting, i.e., under more controllable, standardised conditions than the remote testing used in a large, population-based study (e.g., ^14^).

In each of the affected domains — delayed memory, attention, and executive functioning — between 10 and 20% of patients fell below a cut-off of 1.5 standard deviations based on the healthy group distribution. In fact, a substantial number of patients showed domain-level deficits, as more than half of patients scored below the cut-off in at least one major domain score and just under a fifth of patients were impaired on multiple domains. Deficits were found most commonly in the delayed verbal memory domain. This is in line with the finding of predominant left-sided parahippocampal gyrus atrophy in individuals affected by SARS-CoV-2^25,26^. Interestingly, patients who reported high levels of subjective cognitive complaints exhibited worse performance in the delayed memory domain. As we found no relations between other domains and subjective cognitive complaints, memory deficits may play a unique role in patients’ experience of daily life impairment. We further found relatively high incidences of deficits in attention and executive functioning, which are among the most commonly reported findings in PCS (e.g., ^12-14^).

The use of a large patient sample furthermore allowed for analysing the potential influence of relevant clinical variables on cognitive deficits in patients with PCS. In detail, we tested for the influence of the need for hospitalisation during acute infection, time since infection, relevant comorbidities, and age. Additionally, we tested for the influence of current symptoms of fatigue and depression, which, in line with previous studies (e.g., ^12^, for review see ^27^) were heightened in patients compared to healthy controls.

The analyses revealed, firstly, a — relatively small — negative influence of hospitalisation on memory and attention performance. While reports regarding the effect of disease severity on cognitive functioning in heterogeneous samples of participants following SARS-CoV-2 infection are inconsistent (e.g., ^12-14^), this finding contributes to the understanding of this association with memory and attention in PCS. Secondly, and in accordance with the well-established decline in verbal memory performance with increasing age (for meta-analysis see ^28^), our regression analyses revealed a small influence of age on delayed verbal memory performance. Thirdly, fatigue was a predictor of attention performance, which appears to fit within the context of reduced levels of overall brain arousal and cognitive performance, particularly in the domain of attention (^29,30^, for review see ^31^). As our analyses revealed no associations between cognitive performance and time since infection they suggest that cognitive deficits in the PCS stage may be chronic and no longer improve over time. However, follow-up assessment should provide more conclusive data regarding the long-term course of domain-specific cognitive functioning. Furthermore, as neither depression nor comorbidities were found to be significant predictors, cognitive dysfunctions seem to be due to the infection itself rather than to increased psychological or general health burden.

This study has certain strengths and limitations. Strengths include a large, well-defined post-COVID patient cohort, a socio-demographically matched control group, and the use of an innovative, clinically useful tablet-based assessment tool, which combines resource-efficiency and good psychometric properties. While we did not have access to cognitive performance prior to infection, we mitigated this limitation by including an age- and education-matched control group, as well as by excluding patients with known relevant neurological or psychiatric disorders. Our study was potentially prone to selection bias, as only patients with severe enough symptoms to report to a specialised clinic were included. However, this study thus provides a valuable insight into the clinical cohort, for which the health care system needs to be prepared, as numbers of COVID-19 survivors, who continue to experience long-term symptoms, are rising.

This study identified subtle long-term deficits in attention, memory, and executive functioning persisting for more than three months in patients with PCS. Given the relevance of cognitive deficits for successful reintegration into work and family life, for clinical practice, this indicates a pressing need for the numerous patients suffering from PCS to undergo comprehensive, but time- and cost-efficient cognitive screening, using a tool such as the OCS-Plus, which delivers specific domain-specific information not available from shorter screens. This initial assessment can enable clinicians to decide about further diagnostic and treatment steps, such as the necessity to undergo more in-depth neuropsychological and neurological assessment in specialised centres, or to start treatment with cognitive interventions, such as occupational therapy or computerised training targeting affected domains. From a research perspective, our cross-sectional approach should be complemented by a longitudinal one, i.e., by testing the same patients again in a large-scale follow-up observation.

## Data Availability

All data produced in the present study are available upon reasonable request to the authors.

## Author contributions

VK: Writing – original draft, data curation, formal analysis, data collection. PR: Conceptualisation & implementation, writing – critical review. IU: Data collection. JU: Data collection. ZS: Data collection. ND: Methodology. FR: critical review. MS: critical review. AS: Conceptualisation, supervision, resources, writing – critical review. KF: Conceptualization, supervision, resources, writing – critical review and editing

## Acknowledgments

We would like to thank Claudia Eilert, a representative for the patient support group *Long-COVID Germany* for her expertise and her contributions to the early phases of a project to develop further these mobile cognitive assessment procedures. Additionally, we would like to thank our student assistants Lara Gutfleisch and Antonia Haddenhorst for their contributions to data collection.

## Footnotes

a The present study’s patient sample was recruited from the same outpatient clinic, but using different inclusion criteria (see Methods).

b While the original OCS is a stroke-specific paper-and-pencil bedside screening test, the OCS-Plus is a computerised elaboration of this tool, with broader clinical and subclinical application.

c Due to time constraints in the clinical setting, the OCS-Plus subtask “rule finding” was skipped. Accordingly, scoring for executive functioning differs from the method proposed by Demeyere et al. (2021).

d Effect sizes ranged from 1–.13, which may be classified as small effects^23,24^

