## Supplementary material for "Subtle cognitive impairments in memory, attention, and executive functioning in patients with post-COVID syndrome and their relationships with clinical variables and subjective complaints": Supplemetal Material

**Supplemental material**

S1: Performance on the OCS-Plus domain scales in patients with low versus high complaints of cognitive symptoms

| | Low complaints | | | | | High complaints | | | | | Wilcoxon rank sum test | |
|---|---|---|---|---|---|---|---|---|---|---|---|---|
| **OCS-Plus task** | **n** | **Mean** | **Median** | **SD** | **SEM** | **n** | **Mean** | **Median** | **SD** | **SEM** | **W** | **p** |
| Naming and Semantic Understanding | 78 | 7.87 | 8.00 | 0.37 | 0.04 | 213 | 7.83 | 8.00 | 0.45 | 0.03 | 8599 | 0.224 |
| Memory Encoding | 75 | 9.49 | 10.00 | 0.79 | 0.09 | 207 | 9.43 | 10.00 | 0.73 | 0.05 | 8245.5 | 0.182 |
| Delayed Memory | 75 | 8.35 | 9.00 | 1.62 | 0.19 | 215 | 7.89 | 8.00 | 1.79 | 0.12 | 9214 | 0.03* |
| Praxis | 77 | 102.51 | 105.00 | 12.25 | 1.40 | 213 | 103.14 | 105.00 | 8.32 | 0.57 | 8608.5 | 0.259 |
| Executive Functioning | 75 | 76.87 | 89.86 | 27.97 | 3.23 | 211 | 83.21 | 92.86 | 23.29 | 1.60 | 6889 | 0.959 |
| Attention | 76 | 58.45 | 59.00 | 1.59 | 0.18 | 210 | 58.19 | 59.00 | 1.75 | 0.12 | 8628 | 0.141 |

*Note*. Med. = Median; SD = standard deviation; SEM = standard error of the mean; r = effect size; * = significant before correction; ** = significant after correction

S2: Group-level descriptive statistics for delayed memory, attention, and executive functioning, stratified by presence of comorbidity, within patient group

**Delayed Memory**

| Hypertension | | | | | | Coronary heart disease | | | | | | Chronic heart failure | | | | | | Diabetes mellitus | | | | | | Psychiatric disorders | | | | | |
|---|---|---|---|---|---|---|---|---|---|---|---|---|---|---|---|---|---|---|---|---|---|---|---|---|---|---|---|---|---|
| 0 | | | 1 | | | 0 | | | 1 | | | 0 | | | 1 | | | 0 | | | 1 | | | 0 | | | 1 | | |
| n | M | SD | n | M | SD | n | M | SD | n | M | SD | n | M | SD | n | M | SD | n | M | SD | n | M | SD | n | M | SD | n | M | SD |
| 183 | 8.2 | 1.8 | 96 | 7.6 | 1.8 | 268 | 8 | 1.7 | 11 | 6.9 | 2.6 | 268 | 8 | 1.8 | 11 | 7.6 | 2 | 265 | 8 | 1.8 | 14 | 7 | 1.7 | 239 | 8 | 1.8 | 40 | 8 | 1.7 |

**Attention**

| Hypertension | | | | | | Coronary heart disease | | | | | | Chronic heart failure | | | | | | Diabetes mellitus | | | | | | Psychiatric disorders | | | | | |
|---|---|---|---|---|---|---|---|---|---|---|---|---|---|---|---|---|---|---|---|---|---|---|---|---|---|---|---|---|---|
| 0 | | | 1 | | | 0 | | | 1 | | | 0 | | | 1 | | | 0 | | | 1 | | | 0 | | | 1 | | |
| n | M | SD | n | M | SD | n | M | SD | n | M | SD | n | M | SD | n | M | SD | n | M | SD | n | M | SD | n | M | SD | n | M | SD |
| 181 | 82 | 24.6 | 94 | 80.9 | 25.4 | 264 | 81.8 | 24.6 | 11 | 78 | 30 | 264 | 81.5 | 24.8 | 11 | 82.7 | 25.3 | 261 | 82.1 | 24.3 | 14 | 71.6 | 31.4 | 236 | 82.4 | 24.4 | 39 | 76.4 | 27.1 |

**Executive Functioning**

| Hypertension | | | | | | Coronary heart disease | | | | | | Chronic heart failure | | | | | | Diabetes mellitus | | | | | | Psychiatric disorders | | | | | |
|---|---|---|---|---|---|---|---|---|---|---|---|---|---|---|---|---|---|---|---|---|---|---|---|---|---|---|---|---|---|
| 0 | | | 1 | | | 0 | | | 1 | | | 0 | | | 1 | | | 0 | | | 1 | | | 0 | | | 1 | | |
| n | M | SD | n | M | SD | n | M | SD | n | M | SD | n | M | SD | n | M | SD | n | M | SD | n | M | SD | n | M | SD | n | M | SD |
| 180 | 58.27 | 1.67 | 95 | 58.14 | 1.82 | 264 | 58.25 | 1.72 | 11 | 57.64 | 1.86 | 264 | 58.3 | 1.65 | 11 | 56.45 | 2.5 | 262 | 58.27 | 1.69 | 13 | 57.15 | 2.15 | 236 | 58.27 | 1.71 | 39 | 57.92 | 1.77 |

*Note:* 0 = comorbidity not present; 1 = comorbidity present; n = number of observations per group; M = mean; SD = standard deviation

S3: Wilcoxon Rank Sum Test with Benjamini-Hochberg False Discovery Rate correction for the domains of delayed memory, attention, and executive functioning for each comorbidity

| Domain | Comorbidity | W | p | p_adj |
|---|---|---|---|---|
| Delayed Memory | Hypertension | 10660.5 | 0.003 | 0.045* |
| Executive Functioning | Chronic heart failure | 2126.5 | 0.008 | 0.060 |
| Delayed Memory | Diabetes mellitus | 2482 | 0.030 | 0.150 |
| Executive Functioning | Diabetes mellitus | 2259 | 0.042 | 0.158 |
| Attention | Diabetes mellitus | 2262.5 | 0.117 | 0.340 |
| Executive Functioning | Coronary heart disease | 1786 | 0.186 | 0.340 |
| Delayed Memory | Coronary heart disease | 1811 | 0.191 | 0.340 |
| Executive Functioning | Psychiatric disorders | 5178.5 | 0.200 | 0.340 |
| Attention | Psychiatric disorders | 5162.5 | 0.204 | 0.340 |
| Attention | Coronary heart disease | 1617.5 | 0.505 | 0.678 |
| Attention | Chronic heart failure | 1286.5 | 0.505 | 0.678 |
| Delayed Memory | Chronic heart failure | 1624 | 0.561 | 0.678 |
| Attention | Hypertension | 8832 | 0.588 | 0.678 |
| Executive Functioning | Hypertension | 8785.5 | 0.701 | 0.751 |
| Delayed Memory | Psychiatric disorders | 4735 | 0.924 | 0.924 |

*Note*: p_adj. = FDR-corrected p-value. * = p-value survived correction.

S4: Coefficient-level estimates for models fitted to estimate variation in Delayed Memory performance, taking hypertension into account

| | *Dependent variable:* |
|---|---|
| | Delayed Memory |
| | *OLS* |
| Intercept | $9.542^{***}$ (8.383, 10.789) |
| Hospitalisation (inpatient) | **$-0.701^{**}$ (-1.205, -0.265)** |
| Age | **$-0.024^{*}$ (-0.042, -0.005)** |
| Days since infection | $0.002^{*}$ (0.000, 0.003) |
| FAS | -0.018 (-0.052, 0.013) |
| PHQ-9 | -0.009 (-0.060, 0.042) |
| Hypertension | -0.133 (-0.321, 0.239) |
| Observations | 279 |
| $R^2$ | 0.097 |
| Adjusted $R^2$ | 0.077 |
| Residual Std. Error | 1.701 (df = 272) |
| F Statistic | $4.882^{***}$ (df = 6; 272) |

*Note:* Coefficients and confidence intervals (nonparametric bootstrap, in parentheses), bolded: significant estimates, with bootstrap confidence intervals not overlapping zero; $^{*}p<0.05$; $^{**}p<0.01$; $^{***}p<0.001$
